# Spatial Accessibility Modeling of Vaccine Deserts as Barriers to Controlling SARS-CoV-2

**DOI:** 10.1101/2021.06.09.21252858

**Authors:** Benjamin Rader, Christina M. Astley, Kara Sewalk, Paul L. Delamater, Kathryn Cordiano, Laura Wronski, Jessica Malaty Rivera, Kai Hallberg, Megan F. Pera, Jonathan Cantor, Christopher M. Whaley, Dena M. Bravata, John S. Brownstein

## Abstract

SARS-CoV-2 vaccine distribution is at risk of further propagating the inequities of COVID-19, which in the United States (US) has disproportionately impacted the elderly, people of color, and the medically vulnerable. We identify vaccine deserts - US Census tracts with localized, geographic barriers to vaccine-associated herd immunity - using a comprehensive supply database (VaccineFinder) and an empirically parameterized model of spatial access to essential resources. Incorporating high-resolution COVID-19 burden and time-willing-to-travel for vaccination, we show that early (February – March 2021) vaccine allocation disadvantaged rural and medically vulnerable populations. Data-driven vaccine distribution to vaccine deserts may improve immunization in the hesitant and control SARS-CoV-2.

## Introduction

Geographic access to resources is an essential component to maximizing the benefits of healthcare^1^. Individuals who must travel longer to access services are less likely to receive routine, acute and pandemic-related care^2–4^. To achieve an equitable distribution of resources, spatial barriers should be minimized for all communities and care should be ensured for those who need it the most^1^. This is especially true for the allocation of the SARS-CoV-2 vaccine in the United States (US) given the notable disparities in the virus’s impact^5^. A disproportionate burden of SARS-CoV-2 morbidity and mortality has been experienced among individuals with a lower median income and people of color^6–8^. These differences are associated with significant inequities in the ability to physically distance^9,10^, burden of co-morbidities^7,11^, share of essential workers^7,9,12^, and systemic disparities in the likelihood to die without receiving care^13^. Further, reduced geographic access to SARS-CoV-2 testing in rural areas, areas with a higher percentage of minorities, and areas with a large percentage of uninsured individuals suggests that the disproportionate impact of the virus is likely understated^14^.

The introduction of multiple vaccines creates the potential to reduce the burden of SARS-CoV-2 in the US^15^. Mitigating disparities and ensuring a fair and equitable allocation of the SARS-CoV-2 vaccine are guiding principles of the Centers for Disease Control and Prevention (CDC) distribution recommendations^16^. However, an ideal distribution of the vaccine may be difficult to achieve because it relies on current medical supply chains and distribution locations, which are efficient but demographically biased^14^. The need to vaccinate large numbers of individuals at highly elevated risk for SARS-CoV-2 (e.g. healthcare workers and long-term care residents and staff)^17^ led to initial allocation in most states being focused on hospitals and academic medical centers with supply later being expanded to other clinics and pharmacies. However, these facilities have been shown to be unequally distributed throughout the US and less geographically accessible to rural populations^18–21^. Reliance on these nodes for distribution of the SARS-CoV-2 vaccine can lead to “vaccine deserts”, where local supply is not sufficiently accessible or available.

Studies in Chicago, Baltimore and Los Angeles have shown a higher prevalence of low-access “pharmacy deserts” in minority communities and neighborhoods with higher rates of infectious disease^19,22,23^ while a state-wide study in Pennsylvania showed that pharmacy deserts tend to be in rural communities^20^. Scenario modeling suggests these pharmacy disparities may result in unequal distribution of the SARS-CoV-2 vaccine^24^, but to our knowledge no studies have measured geographic accessibility to confirmed vaccine locations or attempted to account for differences in vaccine supply. In addition, traditional definitions of resource deserts (e.g. food within 10 miles^25^), are insufficient to characterize the unique goals of rapid, population-wide immunization. To quantify accessibility of the SARS-CoV-2 vaccine, we utilize a unique comprehensive dataset of SARS-CoV-2 vaccine providers in the US and employ the enhanced two-step floating catchment area (E2SFCA) method^18^, a technique that has been used to study accessibility to various health resources including primary care physicians^18^, pharmacies^26^, and COVID-19 healthcare^27^. This flexible spatial model simultaneously incorporates supply, demand, travel time, and between-census-tract travel.

In this analysis, we seek to understand if SARS-CoV-2 vaccine doses are being optimally allocated to communities that are most vulnerable to the virus. To accomplish this, vaccine distribution sites and supply measures in the contiguous US states were identified from a comprehensive national database (VaccineFinder.org). Utilizing an E2SFCA model parametrized by people’s willingness to travel to the SARS-CoV-2 vaccine, we then calculated a spatial accessibility score for each US Census tract, the outcome of interest. To connect the relationship of geographic barriers to disease transmission, we define vaccine deserts with a novel modification of resource desert that comports with COVID-19 specific public health goals. Accordingly, we link the E2SFCA accessibility scores to vaccination coverage thresholds that are required to achieve local herd immunity (85%) and evaluate the burden of COVID-19 disparities in the locations that fail to meet these cutoffs (i.e. vaccine deserts) utilizing US Census demographic data and a high-resolution survey of 16.5 million Facebook users. Lastly, to identify if racial and socioeconomic disparities in spatial accessibility to the SARS-CoV-2 vaccine exist within communities of similar population densities, we apply spatial regression modeling and explore a case-series of two highly segregated US cities.

## Data and Methods

### Demographic and Population Data

We gathered demographic and cartographic data from the US Census 2015-2019 American Community Survey^28^ for each census tract in the contiguous US with a non-zero population count (n=72,042, Alaska and Hawaii excluded due to the unique geographic challenges). We calculated population density (people per square kilometer) and the proportional breakdowns by race, sex, median resident age, median income, health insurance type, and internet access. We gathered information about urbanicity (e.g. metropolitan, rural) from the US Department of Agriculture Rural-Urban Commuting Area (RUCA) codes^29^ and population counts from the Gridded Population of the World (v4) dataset^30^.

### COVID-19 and Medical Burden Data

We estimated the COVID-19 and medical burden for each census tract using data from Carnegie Mellon University’s Delphi Group cross-sectional self-reported survey (N=16,533,319) conducted on the Facebook platform (April 6, 2020 – February 11, 2021)^31,32^. Medical burden was quantified using the self-reported proportion of respondents with at least one pre-existing condition that could exacerbate COVID-19 severity (detailed in the supplement). COVID-19 burden was quantified by the proportion of surveys responding “yes” to the question “in the past 24 hours, have you had direct contact with anyone who recently tested positive for COVID-19 (coronavirus)” for each ISO week, which closely mirror government case counts in the US^33,34^, although the latter are only available at county-level resolutions.

### SARS-CoV-2 Vaccine Locations and Supply Data

The location and supply of publicly accessible SARS-CoV-2 vaccine sites in the contiguous US were gathered from VaccineFinder, a comprehensive, national vaccine system maintained by Boston Children’s Hospital in collaboration with the CDC and Castlight Health. VaccineFinder receives registration information and daily reports of on-hand vaccine supply for each SARS-CoV-2 vaccine administration site from the CDC’s Vaccine Tracking System (VTrckS), the ordering platform for entities receiving SARS-CoV-2 vaccines^35^. VTrckS is the most comprehensive vaccine supply source because all 64 public health jurisdictions and pharmacies participating in the Federal Retail Pharmacy Program (FRPP) and receiving direct allocation from the federal government are enrolled^36^. All vaccine distribution sites are collected in the database, however each location can choose to display their location, vaccine availability, and/or appointment resources to the public on the web-based search tool (www.VaccineFinder.org).

We used all vaccine distribution sites (N=37,287) that administered one or more SARS-CoV-2 vaccine dose on at least three separate days between February 17, 2021 – March 16, 2021 (doses administered was imputed from supply levels, details in supplement). The overall dose count for each site was determined by calculating the mean daily doses administered following each site’s receipt of its first reported dose. U.S. Department of Veteran Affairs (VA) locations (extracted from their publicly available database^37^) dispensing the vaccine were also included (N=139). The supply levels for VA vaccine distribution sites were determined by the daily reported difference (February 19, 2021 - March 17, 2021) in cumulative doses administered as reported on February 20, 2021. Dates were chosen based on ready availability of data with the goal of rapid dissemination of research findings.

### Spatial Accessibility Outcome

We calculated spatial accessibility of vaccine supply using the E2SFCA^18^. Scores were undefined for a small number (n=23) of census tracts, resulting in estimates for the majority (n=72,019) of the populated tracts in the contiguous US. The first step of E2SFCA (**Figure S1**, left) calculated the vaccine supply-to-population ratio (*R*_*j*_) for each vaccine distribution site (*j*) based on the mean daily supply (*s*_*j*_) of a site and the population (*P*) of its catchment area (*k* ∈ {*d*_*kj*_ ≤ T}). Daily supply levels were multiplied by 248 to represent the total doses over the aspirational roll out period (January 1 – September 6), assuming the current modeled supply was constant over time. September 6 (Labor Day) was chosen to coincide with typical fall school reopening dates^38^ and the expiration of the majority of COVID-19 related unemployment benefits provided by the American Rescue Plan Act of 2021^39^. These assumptions were chosen to reflect circumstances at the time of analysis. The model used here has the flexibility to incorporate alternative assumptions in the presence of expected change (increases in vaccine supply, etc.).

The catchment populations were calculated using population counts and the travel-time-based weight function (*W*_*d*_) based on willingness to travel to receive the vaccine. We used 1km^2^ resolution gridded data for the population counts and locations in this step^30^. For computational efficiency, we set T equal to 90 minutes.

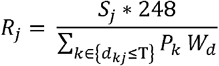

The second step of the E2SFCA (**Figure S1**, right) calculated the spatial accessibility value (*A*) for each census tract (*i*) by summing all the travel-time-weighted supply-to-population ratios *(R*_*j*_*w*_*d*_) for each vaccine distribution site within its own catchment area. Each census tract is represented by its population-weighted centroid^40^. As in the first step, the catchment area is delineated by a travel threshold around each census tract and the supply-to-population ratio is weighted by the travel-time decay function.

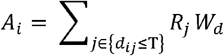

### Travel Time

Travel time (*d*) was determined by utilizing an impedance map of the quickest transversal time (e.g. car, public transportation, boat, etc.) across the gridded population surface of the US^41–43^. For each vaccine distribution location, travel time from all grid squares within 90 minutes were measured by calculating the lowest accumulated travel time across a friction surface using Dijkstra’s algorithm. The same procedure was used to determine the catchment area for each census tract centroid.

### Time Willing to Travel to Vaccine and Travel-Time Decay Function

To empirically define the time unvaccinated individuals would be willing to travel for the SARS-CoV-2 vaccine, we included travel time questions within a large-scale, nationally-representative, previously-validated^44^ web survey from OutbreaksNearMe (ONM) via the SurveyMonkey platform (survey fielded February 9 – February 17, 2021, see reference^45^ for full survey language and **Table S1** for survey demographics). The proportion of respondents previously vaccinated for SARS-CoV-2 was calculated using survey weights. We then asked all unvaccinated survey participants: “What’s the most time you would be willing to travel to receive a COVID-19 vaccine? (please enter a time in minutes).” To ensure comparability to other health resources, we also asked “What’s the most time you would be willing to travel for a routine medical appointment? (please enter a time in minutes).” Responses were limited to 1 to 90 minutes (T in the spatial accessibility model). We received 23,288 responses among those (N=26,466, 88.0%) presented the question. Travel times from survey respondents who indicated they were considering SARS-CoV-2 vaccination (e.g. responded “yes” or “not sure” when asked if they would get the vaccine) were used to define our travel-time decay function^46,47^ (*W*, **Figure 1**, left, plotted as grey line) via a monotonic cubic spline fit to 1 minus the empirical cumulative distribution of these responses. The spline approximates a continuous travel-time delay function (such as the commonly used gaussian function^48^) but captures the different behavioral response to time benchmarks (e.g. perceived difference between 59 minutes and 61 minutes).

**Figure 1:**
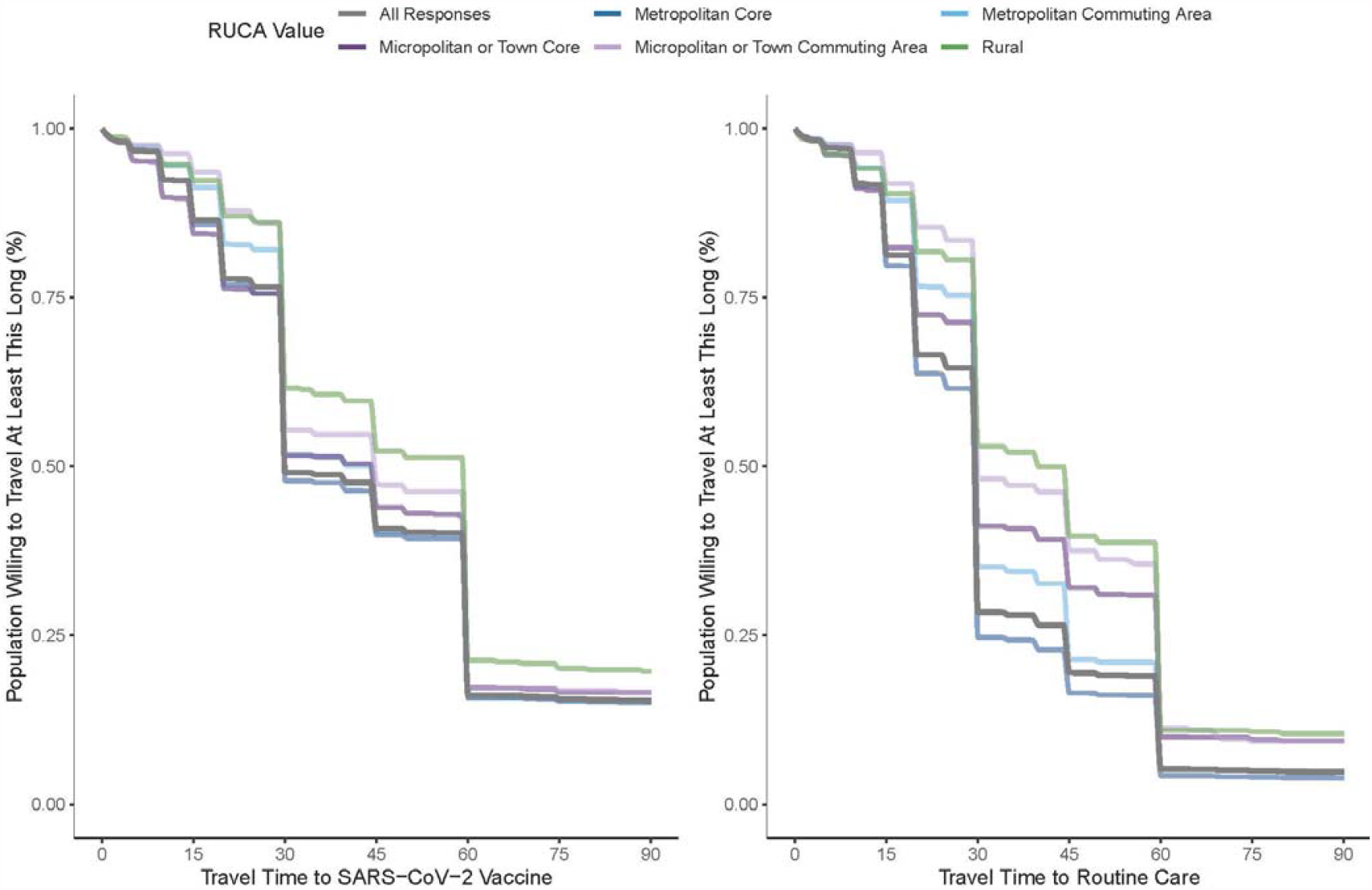
Time Willing to Travel to SARS-CoV-2 Vaccine compared to Routine Healthcare Responses (N=23,288) from US national web survey on how long people were willing to travel to SARS-CoV-2 vaccine (left) compared to routine care (*right*). Lines are monotonic cubic splines fit to 1 minus the empirical cumulative distribution of these response times. Responses were limited to individuals considering a vaccine, defined as those who reported either “yes” or “not sure” when asked if they would get the vaccine. Results are broken down by U.S. Department of Agriculture Rural-Urban Commuting Area (RUCA) codes of the respondent’s self-reported zip code. Overall distribution of travel time to SARS-CoV-2 vaccine (*right, grey*) used as travel-time decay function in E2SFCA model.

### Definition of Vaccine Desert

Access deserts for essential resources (e.g. food, pharmacy) are typically defined in terms of a resource lacking within a specific time or distance^19,23,25^. However, these definitions are restrictive given the lack of consideration of potential supply and our empirical analysis of the long times individuals will travel for a vaccine. Therefore, we used the E2SFCA accessibility score to incorporate supply and travel time into the vaccine distribution scheme model: assuming the vaccine sites available to each census tract have a constant supply for the 8-month period (January – September 6, 2021), each tract would have a travel-time weighted sum of *A*_*i*_ doses per person in their respective weighted catchment areas. We then defined “vaccine deserts” based on a spatial accessibility threshold set in terms of the vaccine access required to achieve herd immunity (assumed 85% of the tract population is vaccinated with one dose, i.e. vaccine desert is *A*_*i*_ < 0.85, additional details in supplement). Similarly, we define low (0.85 ≤ *A*_*i*_ < 1.0), medium (1.0 ≤ *A*_*i*_ < 2.0), and high (*A*_*i*_ ≥ 2.0) accessibility tracts as a function of the population proportion (0.85 or 1) and the number of doses received (1 or 2) of currently available vaccine regimens. Note these thresholds are based on assumptions of the reproductive number, vaccine effectiveness, desired time to achieve herd immunity and a constant supply. Though a simplification, the model allows flexibility to relax these assumptions as additional data become available or if other public health priorities are selected.

### Spatial Regression Model

Based on the known distribution of health resources^49^, we hypothesized that vaccine allocation would be a mixture of consolidated vaccine sites (e.g. medical centers) and intentionally spaced locations to capture markets (e.g. pharmacies, rural health clinics). This may create spatial autocorrelation such that the location of one vaccine distribution site (and resulting accessibility scores) may be dependent on the placement of sites around them. The E2SFCA also allows individuals to access sites across census boundaries, potentially amplifying correlation in accessibility scores when census tracts rely on overlapping distribution points. To quantify the determinants of accessibility accounting for autocorrelation of scores in neighboring census tracts and urban/rural confounding, we fit a spatial lag model:

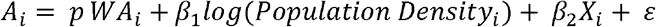

where the term *p wA*_*i*_ represents the coefficient and spatially lagged standardized accessibility score from a weighted matrix (queen contiguity) of neighboring census tracts. Sensitivity analyses of k-nearest (k=4) neighbor inverse distance weighted matrix was performed. We modeled the log of population density and multiple other predictor variables (*X*) for each census tract (*i*) including census demographics and disease burden measures. We then use Monte Carlo simulation to quantify the resulting direct impact^50^, to produce a coefficient similar to Ordinary Least Squares regression and irrespective of spillover effects to neighboring census tracts. Census tracts with incomplete demographic data (N=390, 0.5%) and outlier (see supplement) accessibility scores (N=1,602, 2.2%) were excluded from regression analysis.

Calculations were performed in R version 3.6.2. The ONM survey was approved by the Boston Children’s Hospital Institutional Review Board (IRB-P00023700) and received a waiver of informed consent. The Delphi Group survey protocol was approved by the Carnegie Mellon University IRB (STUDY2020_00000162).

## Results

### Time Willing to Travel Among those Considering Vaccination

In the ONM survey, 19% of survey respondents reported having received at least one dose of SARS-CoV-2 vaccine. The majority of ONM survey respondents (81%) were unvaccinated, and of these, most (N=23,288) indicated “yes” (61.2%) or “not sure” (21.2%) when asked if they wanted the SARS-CoV-2 vaccine. These individuals considering the vaccine indicated they were willing to travel a median of 44.0 [IQR: 30.0-60.0] minutes to receive a SARS-CoV-2 vaccine versus 33.9 [IQR: 20-45] to receive routine medical care (**Figure 1**). Respondents were on average willing to travel 10.6 min (SD 22.3, paired two-sided t-test: p < 0.001) longer to get a vaccine compared to routine care. Notably, while there was a substantial gap between time willing to travel to routine care along an urban-rural divide (31.9 minutes versus 44.0 minutes mean time, respectively, two-sided t-test: t=10.79, p < 0.001) this difference was halved (44.0 minutes versus 50.8 minutes, two-sided t-test: t=5.37, p < 0.001) for travel to the SARS-CoV-2 vaccine. Willingness to travel was more strongly related to vaccine hesitancy (**Figure 2**) than the urban-rural divide, with individuals who were “very eager” to get the vaccine willing to travel a mean of 27.2 minutes longer (two-sided t = 41.1, p <.001) than those who were “very hesitant.”

**Figure 2:**
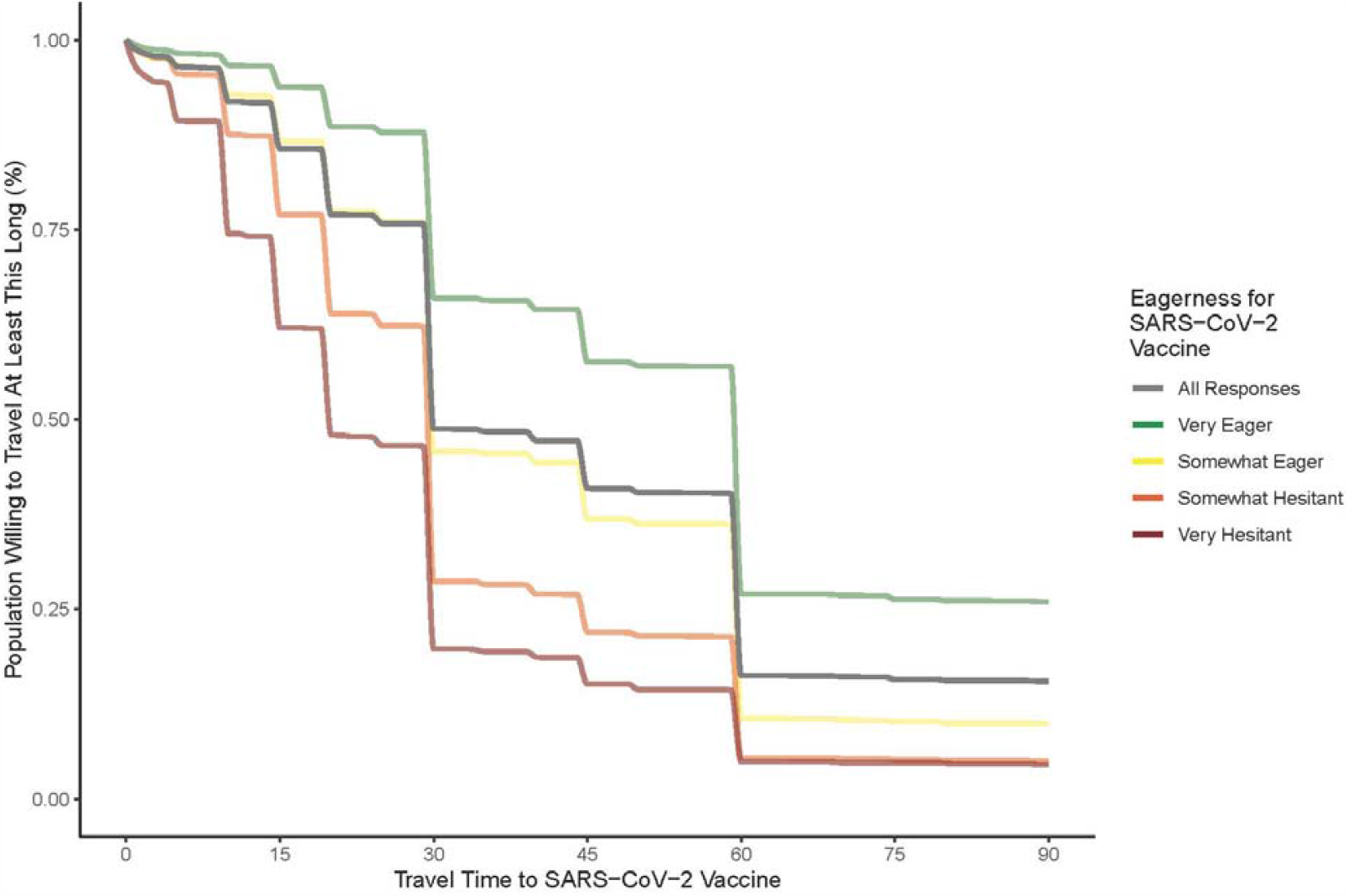
Population Willingness to Travel to SARS-CoV-2 Vaccine by Vaccine Hesitancy Responses Responses (N=23,288) to a large U.S. national web survey on how long people were willing to travel for the SARS-CoV-2 vaccine stratified by self-reported vaccine hesitancy. Lines are monotonic cubic splines fit to 1 minus the empirical cumulative distribution of these response times. Responses were limited to individuals considering a vaccine, defined as those who reported either “yes” or “not sure” when asked if they would get the vaccine.

### Geographic Distribution of Vaccine Deserts

Using the self-reported time-willing-to-travel distribution to parametrize an E2SFCA model of accessibility to the SARS-CoV-2 vaccine sites, we found considerable geographic disparities of vaccine access across the US (**Figure 3)**. There was good accessibility to the vaccine in the Pacific region where over 90% of census tracts were categorized as medium or high accessibility (**Table 1**). Access was also found to be strong in New England and the Middle Atlantic where respectively only 6.5% and 6.3% of census tracts were categorized as vaccine deserts (**Table 1**). Conversely, 32.4% of tracts in the Mountain region were categorized as vaccine deserts as well as 23.4% in West- and 34.1% in East-South Central regions.

**Table 1.** Census Tract Demographics by Accessibility Score for the Contiguous United States

|  | **Vaccine Deserts**<br>N Tracts: 10,912<br>Column Percent | **Low Accessibility**<br>N Tracts: 6,709<br>Column Percent | **Med. Accessibility**<br>N Tracts: 46,735<br>Column Percent | **High Accessibility**<br>N Tracts: 7,663<br>Column Percent | **All**<br>N Tracts: 72,019<br>Row Percent |
| --- | --- | --- | --- | --- | --- |
| **Total Population (N, %)** | 45,699,462 (14.2%) | 31,268,423 (9.7%) | 210,960,831 (65.4%) | 34,483,758 (10.7%) | 322,412,474 (100.0%) |
| **Census Division (N, %)** |  |  |  |  |  |
| New England | 217 (6.5%) | 401 (11.9%) | 1,654 (49.2%) | 1,090 (32.4%) | 3,362 (4.7%) |
| Middle Atlantic | 636 (6.3%) | 619 (6.1%) | 8,123 (80.7%) | 692 (6.9%) | 10,070 (14.0%) |
| East North Central | 1,151 (9.8%) | 816 (7.0%) | 8,310 (70.9%) | 1,436 (12.3%) | 11,713 (16.3%) |
| West North Central | 687 (13.0%) | 396 (7.5%) | 3,503 (66.5%) | 684 (13.0%) | 5,270 (7.3%) |
| South Atlantic | 2,480 (18.3%) | 1,857 (13.7%) | 8,866 (65.4%) | 354 (2.6%) | 13,557 (18.8%) |
| East South Central | 1,514 (34.1%) | 746 (16.8%) | 2,176 (49.1%) | 0 (0.0%) | 4,436 (6.2%) |
| West South Central | 1,895 (23.4%) | 640 (7.9%) | 4,906 (60.6%) | 658 (8.1%) | 8,099 (11.2%) |
| Mountain | 1,692 (32.4%) | 877 (16.8%) | 2,189 (42.0%) | 460 (8.8%) | 5,218 (7.2%) |
| Pacific* | 640 (6.2%) | 357 (3.5%) | 7,008 (68.1%) | 2,289 (22.2%) | 10,294 (14.3%) |
| **Rural-Urban Designation (N, %)** |  |  |  |  |  |
| Metropolitan Core | 3,021 (5.9%) | 3,725 (7.2%) | 37,966 (73.5%) | 6,911 (13.4%) | 51,623 (71.7%) |
| Micropolitan/Town Core | 1,972 (31.3%) | 990 (15.7%) | 2,993 (47.5%) | 344 (5.5%) | 6,299 (8.7%) |
| Metropolitan Commuting Area | 2,463 (33.1%) | 1,132 (15.2%) | 3,588 (48.3%) | 249 (3.4%) | 7,432 (10.3%) |
| Micropolitan/Town Com. Area | 1,830 (52.0%) | 490 (13.9%) | 1,116 (31.7%) | 86 (2.4%) | 3,522 (4.9%) |
| Rural | 1,626 (51.7%) | 372 (11.8%) | 1,072 (34.1%) | 73 (2.3%) | 3,143 (4.4%) |
| **Female (% , Mean, SD)** | 50.0 (±5.0) | 50.6 (±4.9) | 51.0 (±4.7) | 50.8 (±4.2) | 50.8 (±4.8) |
| Missing (N, %) | 20 (0.2%) | 10 (0.1%) | 87 (0.2%) | 13 (0.2%) | 130 (0.2%) |
| **Median Age (Mean Years, SD)** | 42.4 (±8.2) | 40.2 (±8.4) | 39.2 (±7.7) | 38.0 (±6.8) | 39.6 (±7.8) |
| Missing (N, %) | 20 (0.2%) | 14 (0.2%) | 104 (0.2%) | 13 (0.2%) | 151 (0.2%) |
| **Median Income (Mean \$1,000, SD)** | 29.0 (±9.6) | 31.9 (±11.8) | 33.7 (±14.1) | 37.5 (±17.1) | 33.2 (±13.9) |
| Missing (N, %) | 32 (0.3%) | 19 (0.3%) | 144 (0.3%) | 18 (0.2%) | 213 (0.3%) |
| **Race/Ethnicity (Mean %, SD)** |  |  |  |  |  |
| White | 80.1 (±21.1) | 77.7 (±22.7) | 70.5 (±25.7) | 67.2 (±25.2) | 72.3 (±25.1) |
| Black | 10.5 (±17.9) | 12.6 (±20.4) | 15.1 (±22.8) | 12.8 (±21.0) | 13.9 (±21.8) |
| Hispanic | 10.9 (±16.1) | 13.6 (±18.4) | 18.4 (±22.8) | 17.9 (±20.8) | 16.7 (±21.5) |
| American Indian or Alaskan Native* | 2.2 (±9.6) | 0.9 (±3.6) | 0.6 (±2.7) | 0.6 (±2.2) | 0.9 (±4.6) |
| Asian | 1.7 (±3.5) | 2.5 (±4.6) | 5.2 (±9.0) | 9.4 (±13.5) | 4.9 (±8.9) |
| Hawaiian or Pacific Islander* | 0.1 (±0.5) | 0.1 (±0.4) | 0.1 (±0.6) | 0.2 (±0.9) | 0.1 (±0.6) |
| Two or More Races | 2.7 (±2.8) | 2.8 (±2.6) | 3.2 (±2.7) | 4.0 (±3.3) | 3.1 (±2.8) |
| Other Race | 2.6 (±5.9) | 3.3 (±6.4) | 5.2 (±9.4) | 5.7 (±9.1) | 4.7 (±8.7) |
| Missing (N, %) | 20 (0.2%) | 10 (0.1%) | 87 (0.2%) | 13 (0.2%) | 130 (0.2%) |
| **No Internet Access (Mean %, SD)** | 19.8 (±11.4) | 15.6 (±10.2) | 14.4 (±10.2) | 13.1 (±9.7) | 15.2 (±10.6) |
| Missing (N, %) | 43 (0.4%) | 30 (0.4%) | 209 (0.4%) | 22 (0.3%) | 304 (0.4%) |
| **Insurance Coverage (Mean %, SD)** |  |  |  |  |  |
| Employer | 40.7 (±12.9) | 44.5 (±13.7) | 45.5 (±15.2) | 49.1 (±15.3) | 45.1 (±14.9) |
| Direct | 6.4 (±4.0) | 6.4 (±4.0) | 6.6 (±4.3) | 6.7 (±4.2) | 6.5 (±4.2) |
| Medicare | 6.6 (±3.4) | 5.7 (±3.5) | 5.3 (±3.1) | 4.7 (±2.6) | 5.5 (±3.2) |
| Medicaid | 15.0 (±9.4) | 14.6 (±10.5) | 16.1 (±12.7) | 16.4 (±12.9) | 15.8 (±12.1) |
| Veterans Administration | 0.4 (±0.5) | 0.3 (±0.5) | 0.3 (±0.5) | 0.2 (±0.5) | 0.3 (±0.5) |
| Tricare | 1.2 (±4.4) | 1.3 (±4.9) | 0.9 (±3.7) | 0.5 (±1.9) | 0.9 (±3.8) |
| Two or More | 6.5 (±3.3) | 6.4 (±3.3) | 5.9 (±3.2) | 5.8 (±3.1) | 6.0 (±3.2) |
| Uninsured | 10.1 (±6.6) | 9.1 (±6.6) | 8.8 (±7.2) | 6.8 (±5.6) | 8.8 (±7.0) |
| Missing (N, %) | 38 (0.3%) | 23 (0.3%) | 153 (0.3%) | 16 (0.2%) | 230 (0.3%) |
| **Medical Burden<sup>†</sup> (Mean %, SD)** | 53.4 (±5.8) | 51.4 (±5.6) | 48.4 (±6.3) | 46.3 (±6.4) | 49.2 (±6.5) |
| Missing (N, %) | 72 (0.7%) | 15 (0.2%) | 76 (0.2%) | 4 (0.1%) | 167 (0.2%) |
| **COVID-19 Burden<sup>†</sup> (Mean %, SD)** | 3.1 (±1.3) | 2.8 (±1.1) | 2.6 (±1.1) | 2.2 (±1.1) | 2.7 (±1.1) |
| Missing (N, %) | 72 (0.7%) | 15 (0.2%) | 76 (0.2%) | 4 (0.1%) | 167 (0.2%) |

\*Alaska and Hawaii excluded from analysis <sup>†</sup>Percentage of survey respondents in each census tract who reported COVID-19 related pre-existing conditions (medical burden) or previous contact with COVID-19 positive individual (COVID-19 burden).

**Figure 3:**
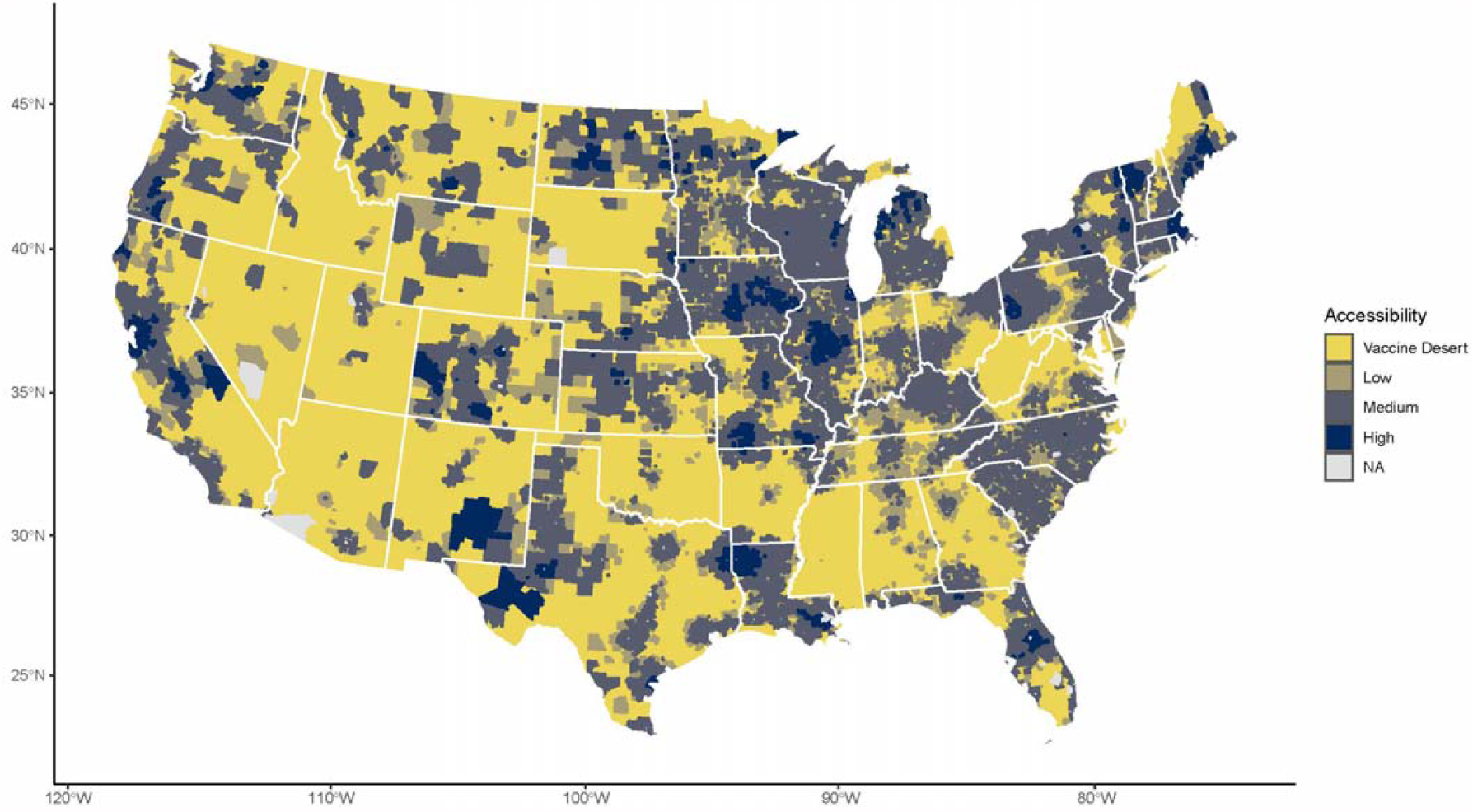
SARS-CoV-2 Vaccine Deserts and Accessibility Scores Across the United States Map of United States Census tracts colored by their spatial accessibility to SARS-CoV-2 vaccine doses. Accessibility scores (*A*_*i*_) were estimated by the enhanced two-step floating catchment area method. Access strata broadly represent cutoffs suggesting difficulty in reaching herd immunity in 8-months (Jan 1 – American Labor Day) with one dose (vaccine desert, *A*_*i*_ < 0.85) immunizing all with one dose (low, 0.85 ≤ *A*_*i*_ < 1.0), difficulty immunizing all with two doses (medium, 1.0 85 ≤ *A*_*i*_ < 2.0), and sufficient accessibility (high, *A*_*i*_ ≥ 2.0). Under the modeled distribution scheme, if the vaccine sites available to each census tract have a steady supply for the rollout period, they would have a sum (accounting for travel-time decay) of *A*_*i*_ doses per person in their respective weighted catchment areas. No estimates (NA) were provided for non-populated census tracts and those (n=23) where *A*_*i*_ was undefined.

### Vaccine Deserts in Relation to Demography

Vaccine deserts are home to 14.2% (N=45,699,462) of the US census estimated population. When stratifying census tracts by RUCA code, spatial accessibility is higher in metropolitan cores (86.9% medium or high accessibility). This falls dramatically with RUCA classifications farther from metropolitan cores, dropping from 53.0% (micropolitan/town core) to 36.4% (rural area). Rural and micropolitan/town commuting areas had the worst accessibility; vaccine deserts comprised 42.9% and 41.3% of census tracts in these areas, respectively.

In contrast to other forms of vaccine access, disparities in spatial access largely reflect the demographics of the urban-rural divide. Vaccine deserts as compared to high accessibility locations are concentrated among older (4.4 yr. more), white (13.0% more), lower income ($8,579 less), and uninsured (3.4% higher) census tract populations. Vaccine deserts also have a greater burden of those with COVID-19-associated medical conditions (7.1% higher), COVID-19 exposure (0.8% higher), and less access to the internet (6.7% less). These relationships scale monotonically, and low and medium accessibility areas have differences that fall linearly between vaccine deserts and high accessibility locations.

### Determinants of Spatial Accessibility

To understand the determinates of spatial accessibility accounting for urban-rural confounding, we performed separate spatial regression analyses for each variable of interest while controlling for population density. Census tracts in the highest quartile of Black residents and Hispanic residents were associated with decreased accessibility to the SARS-CoV-2 vaccine when compared to the lowest quartile for each variable (**Table 2**). Tracts with older residents and a higher prior COVID-19 burden were also associated with decreased accessibility. We find the opposite effect for medical burden whereby, when controlling for population density, an increased percentage of individuals with self-reported pre-existing conditions was associated with higher access to the SARS-CoV-2 vaccine. All results were robust to a sensitivity analysis with an alternative weighting matrix (**Table S2**).

**Table 2.** Spatial Regression on Accessibility to SARS-CoV-2 Vaccine Score (*A*_*i*_) for each US Census Tract

|  | Model 1: Population Density |  | Model 2: Percent Black |  | Model 3: Percent Hispanic |  | Model 4: Age |  | Model 5: Medical Burden<sup>‡</sup> |  | Model 6: COVID-19 Burden<sup>‡</sup> |  |
| --- | --- | --- | --- | --- | --- | --- | --- | --- | --- | --- | --- | --- |
|  | Coefficient | [Direct<sup>†</sup>] | Coefficient | [Direct<sup>†</sup>] | Coefficient | [Direct<sup>†</sup>] | Coefficient | [Direct<sup>†</sup>] | Coefficient | [Direct<sup>†</sup>] | Coefficient | [Direct<sup>†</sup>] |
| **Log of Population Density** | 0.013*** | [0.019] | 0.014*** | [0.020] | 0.014*** | [0.020] | 0.012*** | [0.018] | 0.014*** | [0.020] | 0.013*** | [0.019] |
| **Percent Black Race** |  |  |  |  |  |  |  |  |  |  |  |  |
| Q1 (0% – 1.0%] |  |  | REF | REF |  |  |  |  |  |  |  |  |
| Q2 (1.0% – 4.3%] |  |  | -0.001** | [-0.001] |  |  |  |  |  |  |  |  |
| Q3 (4.3% – 15.6%] |  |  | -0.009*** | [-0.013] |  |  |  |  |  |  |  |  |
| Q4 (15.6% – 100%] |  |  | -0.010*** | [-0.017] |  |  |  |  |  |  |  |  |
| **Percent Hispanic Ethnicity** |  |  |  |  |  |  |  |  |  |  |  |  |
| Q1 (0% – 2.8%] |  |  |  |  | REF | REF |  |  |  |  |  |  |
| Q2 (2.8% – 7.6%] |  |  |  |  | 0.002 | [0.003] |  |  |  |  |  |  |
| Q3 (7.6% – 20.8%] |  |  |  |  | -0.006*** | [-0.008] |  |  |  |  |  |  |
| Q4 (20.8% – 100%] |  |  |  |  | -0.007*** | [-0.009] |  |  |  |  |  |  |
| **Median Age (years)** |  |  |  |  |  |  | -0.001*** | [-0.001] |  |  |  |  |
| **Medical Burden (%)** |  |  |  |  |  |  |  |  | 0.040*** | [0.058] |  |  |
| **COVID-19 Burden (%)** |  |  |  |  |  |  |  |  |  |  | -0.276*** | [-0.394] |
| **Spatial Lag ($\rho$)** | 0.923*** |  | 0.922*** |  | 0.922*** |  | 0.923*** |  | 0.923*** |  | 0.922*** |  |
| **N (Census Tract)** | 70027 |  | 70027 |  | 70027 |  | 70027 |  | 70027 |  | 70027 |  |
| **AIC** | -66245 |  | -66363 |  | -66310 |  | -66408 |  | -66296 |  | -66375 |  |

\*p < 0.05, \*\*p < 0.01, \*\*\*p < 0.001
<sup>†</sup>Direct impact can be interpreted similar to traditional OLS regression coefficient
<sup>‡</sup>Percent of survey respondents in each census tract who reported COVID-19 related pre-existing conditions (medical burden) or previous contact with COVID-19 positive individual (COVID-19 burden).

### Case Study of Racial Disparities in Highly Segregated Urban Centers

To explore differential access by race within urban areas, we focus on two highly racially segregated cities, Detroit (defined here: Wayne, Oakland, and Macomb Counties, MI) and Chicago (Cook County, IL)^51^. In both cities, a qualitative analysis shows census tracts with a higher proportion of Black residents are served by noticeably fewer vaccine distribution sites compared to nearby tracts with fewer Black residents (**Figure 4**). Of note, vaccine distribution sites located in minority neighborhoods in Detroit are more centralized consisting of fewer locations with larger supply, while tracts with a larger percentage of white residents are served by a distributed network of smaller sites. In Chicago we see similar disparities. In the South Side of the Chicago and the neighboring areas of Cook County, we see far fewer vaccination sites than in the North and Near North parts of the city. We also found that the largest distribution sites in Chicago are all located in predominately non-Black race regions of the city.

**Figure 4:**
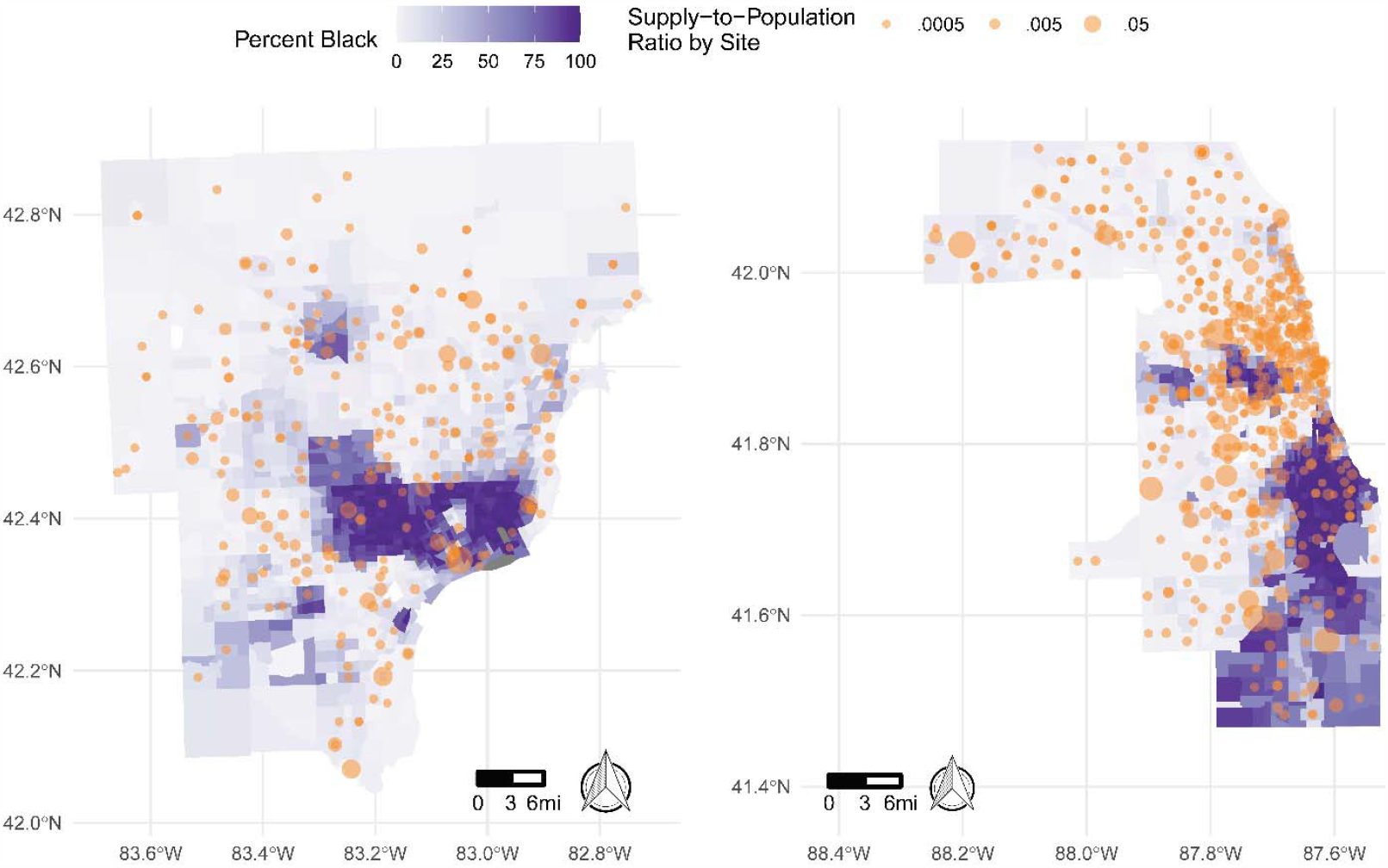
Disparities in SARS-CoV-2 Vaccine Access in Detroit and Chicago, USA Map of census tracts around Detroit, Michigan (*left*) and Chicago, Illinois (*right*) by the percentage of residents who are Black (*purple*). Overlayed in orange are SARS-CoV-2 distribution locations sized by their supply-to-population ratio. Vaccine distribution sites in both cities are sparser in census tracts with a higher percentage of Black residents.

## Discussion

The distribution of the SARS-CoV-2 vaccine supply during the timeframe of February 19, 2021 - March 17, 2021 has created vaccine deserts that have the potential to interfere with population-wide disease control. Based on an empirically-parameterized spatial accessibility model, paired with an extensive national dataset of confirmed locations with vaccines and their doses administered, we find that the present allocation scheme favors spatial accessibility within US metropolitan areas. In contrast, vaccine deserts, more often found in rural regions, have a preponderance of vulnerable populations. Vaccine deserts have more residents with self-reported COVID-19 exposures and pre-existing conditions as well as more individuals who lack health insurance. When measuring determinates of accessibility while controlling for population density, we see that tracts with a higher proportion of Black residents, Hispanic residents and older residents have reduced spatial access to the vaccine. Together, these findings highlight that at the time of our analysis (Feb 19 - March 17 2021) vaccine dissemination points within the US were not yet effectively placed to target those most vulnerable to SARS-CoV-2.

US populations living in rural census tracts have the lowest accessibility to the SARS-CoV-2 vaccine. While prior studies hypothesize that rural populations may be willing to travel further distances for health resources than those in urban areas^52,53^ we find these differences are diminished for the SARS-CoV-2 vaccine. Despite a large portion of urban populations’ willingness to travel longer for the vaccine than they would for routine care, this effect is abated for those in rural census tracts. This discrepancy may be fueled by rural populations reporting higher levels of vaccine hesitancy^54^, which our findings show is a stronger predictor of willingness to travel than geography. These regions may benefit the most from the SARS-CoV-2 vaccine due to their relatively higher medical burden^55^, older population^56^ and lower likelihood to consider adoption of mitigation measures such as face mask wearing^44^. We also find that convenience could play a large role in whether hesitant individuals would be willing to get vaccinated, suggesting increased spatial access may be a meaningful public health intervention. Only approximately 50% of the overall population reports willingness to travel more than 30 minutes for the vaccine and less than 20% would travel more than one hour. These numbers are even smaller amongst hesitant individuals. Despite rural areas being less populated than urban ones, after incorporating these findings, our model suggests approximately 14% of the US lives in a vaccine desert at the time of analysis. Inability to reach spatially coupled rural regions with vaccinations presents the risk of intense outbreaks^57^ and may sustain persistent chains of transmission which are barriers to infection elimination^58^, although the latter has yet to be studied for SARS-CoV-2.

SARS-CoV-2 morbidity and mortality is not distributed equally across demographic groups and prior work has shown that within both urban^9,13^ and rural^59^ regions, communities with a higher proportion of minorities had worse outcomes. Ensuring equal access to the SARS-CoV-2 vaccine is an important part of mitigating structural racism and preventing further propagation of these imbalances. Previous statewide analyses have shown that pharmacy deserts are more likely to be in rural regions with a high white population^20^, while citywide analyses have shown they tend to be located in neighborhoods with more minorities^19,22,23^. Utilizing a 90-minute catchment area to understand vaccine deserts (the definition of which is specific to COVID-19 immunization goals, but more flexible than the strict radius limits routinely used to study access to less scarce resources^19,23^), our present results are congruent with both these findings. On the national level, we observe vaccine deserts in rural census tracts that are predominately white.

Yet, when we remove urban-rural confounding and look within regions of similar spatial structure, we find the same racial disparities highlighted in the pharmacy desert literature, suggesting current vaccine allocation strategies do not go far enough to address these disparities. Conversely, when adjusting for urban-rural confounding we do find higher spatial accessibility for those with self-reported pre-existing conditions, which may highlight deliberate vaccine site placement or the likelihood of individuals who live near medical resources to receive diagnoses^60^. While self-reports are biased estimates of medical diagnoses, this also suggests that our general finding of higher burden in rural areas may be an underestimate given a rural region’s reduced access to diagnoses.

Importantly, the geospatial barriers discussed here only represent one type of vaccine access challenge. Social disparities (e.g. ability to take time off work, obtain child care, procure efficient transportation) suggest equal spatial barriers can manifest very differently^61,62^ and that vulnerable populations may be disproportionately affected by poor spatial access. For example, this is supported by evidence suggesting that individuals with lower income experienced a disproportionate reduction in access to health services in response to the pandemic^63^. The two-dose regimens for the selected SARS-CoV-2 vaccine formulations available at the time of analysis also create a need to access a vaccine distribution site twice within a short period (21 or 28 days), suggesting that spatial barriers may compound and/or affect dose completion rates. Additionally, the disparities in internet access in vaccine deserts suggest these regions may have a harder time both identifying and accessing vaccine distribution locations. These same disparities in internet access may affect the representativeness of the two surveys used here and cause undersampling in vaccine deserts (which census data shows have poorer internet access). Therefore, our results may underestimate the medical and COVID-19 burden in census tracts with poor spatial accessibility to the SARS-CoV-2 vaccine.

The VaccineFinder dataset used in this analysis is likely the most comprehensive source of vaccine site and dose supply information given the mandate for SARS-CoV-2 vaccine providers enrolled in the FRPP to register in VTrckS. However, not all locations modeled may be open or advertising to the public and we only included locations with evidence of dose administration on multiple occasions. These locations will likely change substantially as vaccine supply expands to more providers. Additionally, while we integrate locations from the Veterans Administration, our dataset is missing sites in states that only partially joined FRPP (e.g. West Virginia) and other important federal vaccine distribution channels such as Military Treatment Facilities, the Indian Health Service and Federally Qualified Health centers.

The current analysis also only measured spatial access from the perspective of the dose recipient (and assumed everyone in the population was eligible to receive a vaccine) based on static observations of vaccination locations in the early phases of the rollout process. Distribution during this period was characterized by manufacturing constraints^17^ and supply chain barriers (e.g. need for ultralow temperature freezers)^64^, as well as prioritized vaccination of healthcare workers, those over 65 years of age, and high-risk individuals^17^. The selected site locations during the time of this analysis favor urban areas to optimally reach these populations for vaccination. Populations with a high number of medical workers may be vaccinated relatively quickly, allowing some distribution nodes to broaden their population served. Conversely, populations with a high number of children under 18 have a large proportion of residents who may not yet be eligible for the vaccine. Additionally, this analysis did not account for natural or possible vaccine-derived immunity levels already in the population which is rapidly changing and may not be evenly geographically distributed.

As vaccine priorities evolve, states increase/shift distribution locations and supply, and one-dose vaccines are integrated into the supply-chain, the barriers identified here can be overcome and the current results will change. This manuscript presents a novel utilization of a highly flexible model that can incorporate these future changes while also helping identify the need to direct vaccine supply to undeserved communities. Controlling COVID-19 in the US through herd immunity requires extensive, well-orchestrated coordination of vaccine delivery at the national and local scales^65^. Here we show significant disparities in geographic access to the SARS-CoV-2 vaccine during the early stages of the vaccination program, a hurdle to this achievement.

Eliminating SARS-CoV-2 vaccine deserts and spatial barriers to transmission control will require an ongoing commitment to optimizing distribution to align with continued expansion of populations prioritized to receive vaccines and simultaneous investment in resources to mitigate transportation access disparities. This will necessitate locally targeted interventions including active outreach campaigns, mobile vaccination clinics and other innovative health delivery mechanisms such as “vaxathons^66^” and ride-share partnerships^67^.

## Data Availability

Real-time vaccine site location is available at VaccineFinder.org for locations that choose to publicly display. Raw data is not available for sharing. Delphi Group Facebook COVID-19 Symptom Survey data may requested via [https://cmu-delphi.github.io/delphi-epidata/].

## Code Availability

Code and travel-time decay function will be available: github.com/computationalepi/E2SFCA.

## Acknowledgements

This work was supported by Flu Lab and Ending Pandemics. BR, CMA and JSB acknowledge funding from Facebook Sponsored Research Agreement [INB1116217]. BR and JSB acknowledge funding from Google.org via the Tides Foundation [TF2003–089662]. The authors thank Kate Miller and Ethan Roubenoff for their insights and assistance. The authors thank the HealthMap VaccineFinder and research teams including Christopher Remmel, Gaurav Tuli and Autumn Gertz. The authors thank the SurveyMonkey research team including Jon Cohen and Jack Chen. The authors also thank the Castlight Health team including Vivek Salgar, Ankur Jain, Rajiv Kutty, Mukul Mittal, Gavin Thompson, Sophia Golden, Danielle Hershon, Mustafa Husaini, Ariel Klein, Venkatesh Hemanth Gorur, Kranthi Ravi, Akshay Mungrey, Terry Bennett, Phani Kumar Mallampati, Vaneet Loomba, Abhishek Jain, Vamshi Krishna Naidi, Umamaheshwara Gupta Karnati, Stephen Brokaw, Brian Quock, Deepak Mali, Milind Pandrangi, Ryan Scribner and Shahram Moatazedi. Boston Children’s Hospital’s High-Performance Computing Cluster Enkefalos 2 (E2) was used for research reported in this publication. Software used in the project was installed and configured by BioGrids^68^.

## Contributions

BR, CMA, DMB and JSB conceived of the study. BR, CMA, KS, KC and JSB wrote the first draft of the manuscript. KS, LW, KH, DMB and JSB oversaw and implemented data collection. BR, CMA, PLD and KH contributed to analysis. All authors contributed to the interpretation of the data and critical revision of the final manuscript. BR and JSB have verified the underlying data. All authors have seen and approve the final text.

