## Supplement for "Spatial Accessibility Modeling of Vaccine Deserts as Barriers to Controlling SARS-CoV-2"

**Doses Administration Days**

The VaccineFinder database contains the number of doses each site reported per day. Doses administered was measured as the difference between doses each day and the day prior. Days with negative values (e.g. more doses on Tuesday than Monday) were considered “supply days” and excluded from analysis. Only locations with three or more dose administration days were included. This method allowed for the systematic exclusion of centralized warehouses reporting large dose quantities, but not administering doses to individuals. This approach may lead to underestimating total dose quantity administered in exchange for better capturing precise locations where doses are being administered.

**Delphi Survey**

*List of Pre-Existing Conditions:*

1. Diabetes
2. Cancer (other than skin cancer)
3. Heart disease
4. High blood pressure
5. Asthma
6. Chronic lung disease such as COPD or emphysema
7. Kidney Disease
8. Autoimmune disorder such as rheumatoid arthritis or Crohn’s disease

*Self-reported Percent of U.S. with Pre-existing Conditions (numbers from list above):*

| **1** | **2** | **3** | **4** | **5** | **6** | **7** | **8** | **None** | **N** |
| --- | --- | --- | --- | --- | --- | --- | --- | --- | --- |
| 10.9% | 5.0% | 5.3% | 28.0% | 15.2% | 4.1% | 2.1% | 7.4% | 46.8% | 16,533,319 |

*Disease Burden Validation*

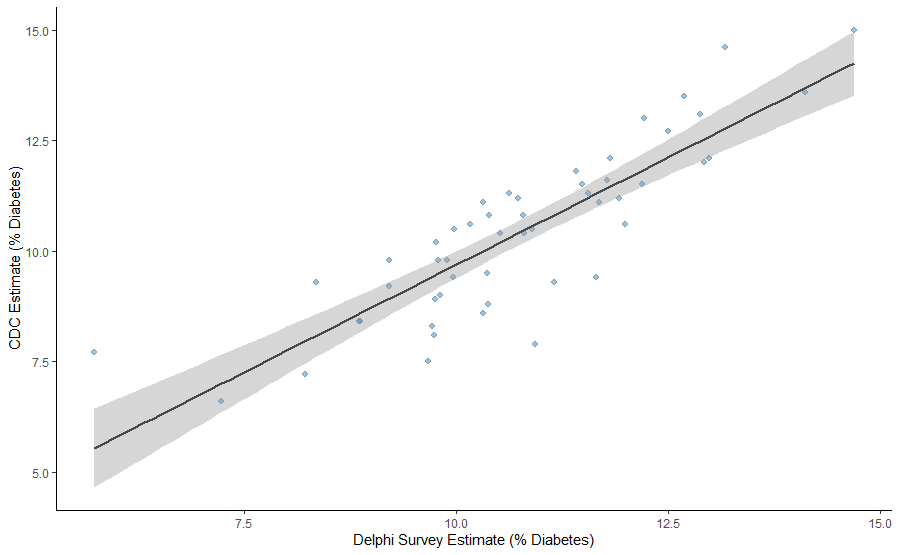
We compared estimates of diabetes from the Delphi survey to CDC estimates of Adults Diagnosed with diabetes^1^ for each of the 50 states and the district of Columbia (Pearson’s *r* = .86):

*Zip to Census Tract Mapping and Validation*

Zip codes were mapped to census tracts such that each individual survey response contributed to all census tracts it overlapped with, proportional to the percentage of overlap (as measured by residential addresses)^2,3^. For example, a survey response from zip 40025 contributed 0.99 persons to census tract 21111007502 and 0.01 persons to census tract 21111010318 because this zip code had a 99% and 1% overlap with the census tracts, respectively. Census tracts containing under 10.0 total persons were excluded from analyses.

The goal of this sub-analysis is to understand how accessibility relates to comorbidities and COVID-19 burden at the census tract level. However, some variables such as COVID-19 burden are only measured at the county and zip code level as opposed to the census tract. Of primary concern is whether zip code data mapped to census tract in the above-described method (hence “derived census tract”) is an adequate representation of what is happening at the census tract level. An alternative is to use lower resolution county data as a stand-in for census tract variables.

Given that the average age of residents is available at all three spatial resolutions, we compare correlations in this known variable to understand which method may be best to represent variables unmeasured in census tracts (e.g. COVID burden). First, we compare percentage of survey respondents over age 65 in each derived census tract with ACS estimates of individuals over 65 in each census tract (Pearson’s *r* = .65). Next, we compare the percentage of individuals over 65 in each census tract to the same variable inherited from the county estimate (Pearson’s *r* = .49). While neither is a perfect representation of what is happening in the census tract, utilizing derived census tracts explains more variation in actual census tract age than using estimates inherited from a county resolution.

**Case Study City Boundaries**

Detroit was defined by Wayne, Oakland and Macomb County borders. Chicago was defined by Cook County borders.

**Census Tracts Outliers**

Outliers were census tracts with accessibility scores greater than 2 standard deviations over the mean. Qualitatively, outliers were generally remote census tracts, adequately served by a single vaccine location, with little competition for doses within 90 minutes.

**Herd Immunity Estimates**

$$V_{c_{1}}= \frac{1}{Ve_{1}}*(1- \frac{1}{R_{0}})$$

One dose vaccine coverage ($V_{c_{1}}$) is estimated to be approximately 85% as a function of:

- The approximate mean first dose vaccine efficacy ($Ve_{1}$) of the BNT162b2 and mRNA-1273 (80%) vaccines following appropriate waiting period^4^
- A mean estimated basic reproductive number ($R_{0}$) of SARS-CoV-2 (3.28)^5^
- Assuming random mixing and persistent immunity
- We did not include pre-existing infection derived immunity

**Supplement References:**

^1^CDC Diabetes Atlas: https://gis.cdc.gov/grasp/diabetes/DiabetesAtlas.html

^2^Cross-walk files: [https://www.huduser.gov/portal/datasets/usps_crosswalk.html](about:blank)

^3^Din, A. & Wilson, R. Crosswalking ZIP Codes to Census Geographies. Cityscape 22, 293–314 (2020).

^4^Thompson, M. G. et al. Interim Estimates of Vaccine Effectiveness of BNT162b2 and mRNA-1273 COVID-19 Vaccines in Preventing SARS-CoV-2 Infection Among Health Care Personnel, First Responders, and Other Essential and Frontline Workers — Eight U.S. Locations, December 2020–March 2021. MMWR. Morb. Mortal. Wkly. Rep. 70, 495–500 (2021).

^5^Liu, Y., Gayle, A. A., Wilder-Smith, A. & Rocklöv, J. The reproductive number of COVID-19 is higher compared to SARS coronavirus. J. Travel Med. 27, 1–4 (2020).

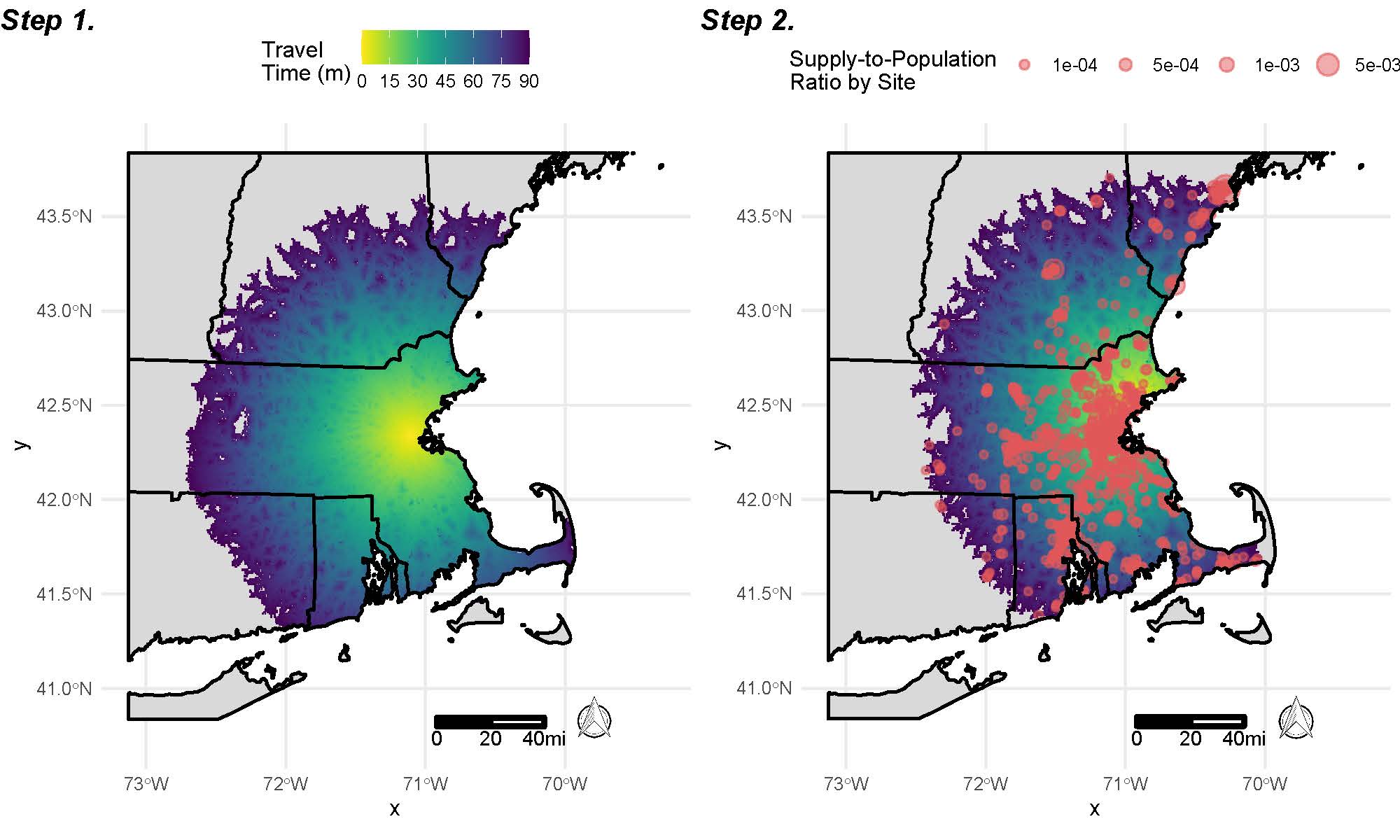
**Figure S1:** Example of Step One and Two of Enhanced Two Step Catchment Area Method from Massachusetts and Surrounding States

The first step of E2SFCA is to determine the supply-to-population ratio of each vaccine provider. The catchment area (the denominator of this equation) is defined as the population in each 1km grid square within 90 minutes of the site, weighted by the travel time from the grid square to the site (e.g. each site can serve 100% the population within 1 minute of the site and 50% of the population 40 minutes away). The numerator of the equation is determined by the daily doses on hand for each site. This process is repeated for each distribution location (*left*, shows example from Boston Children’s Hospital Site). The second step is to calculate accessibility scores of each census tract by taking the sum of all the supply-to-population ratios (*salmon*) from within 90 minutes of the location. The supply-to-population ratios from sites further away are similarly downweighed. This step is repeated for each census tract in the United Sates (*right*, shows example from census tract in Salem, MA).

**Table S1** OutbreaksNearMe/SurveyMonkey Respondent* Demographics by Minutes-Willing-to-Travel to SARS-CoV-2 Vaccine

|  | **(1,15]** | **(15,30]** | **(30,45]** | **(45,60]** | **(60,90]** | **Total** |
| --- | --- | --- | --- | --- | --- | --- |
|  | **No. 3,702** | **No. 10,139** | **No. 2,171** | **No. 7,001** | **No. 4,381** | **No. 27,394** |
| Age | | | | | | |
| Mean (SD) | 48.9 (±16.4) | 52.8 (±16.3) | 53.1 (±17.1) | 53.8 (±16.2) | 52.1 (±16.7) | 52.4 (±16.5) |
| Missing | 341 (9.2%) | 715 (7.1%) | 119 (5.5%) | 392 (5.6%) | 246 (5.6%) | 1,813 (6.6%) |
| Gender | | | | | | |
| Female | 2,102 (13.4%) | 5,997 (38.1%) | 1,247 (7.9%) | 4,021 (25.6%) | 2,362 (15.0%) | 15,729 (57.4%) |
| Male | 1,185 (12.5%) | 3,307 (34.8%) | 782 (8.2%) | 2,521 (26.5%) | 1,709 (18.0%) | 9,504 (34.7%) |
| Other | 74 (21.3%) | 120 (34.5%) | 23 (6.6%) | 67 (19.3%) | 64 (18.4%) | 348 (1.3%) |
| Missing | 341 (18.8%) | 715 (39.4%) | 119 (6.6%) | 392 (21.6%) | 246 (13.6%) | 1,813 (6.6%) |
| Race/Ethnicity | | | | | | |
| White | 1,981 (10.9%) | 6,452 (35.5%) | 1,505 (8.3%) | 4,967 (27.3%) | 3,265 (18.0%) | 18,170 (66.3%) |
| Black | 558 (20.4%) | 1,181 (43.1%) | 201 (7.3%) | 537 (19.6%) | 265 (9.7%) | 2,742 (10.0%) |
| Hispanic | 332 (16.7%) | 772 (38.8%) | 155 (7.8%) | 473 (23.7%) | 260 (13.1%) | 1,992 (7.3%) |
| Asian | 235 (17.2%) | 522 (38.2%) | 101 (7.4%) | 345 (25.3%) | 163 (11.9%) | 1,366 (5.0%) |
| Other | 255 (19.5%) | 497 (37.9%) | 90 (6.9%) | 287 (21.9%) | 182 (13.9%) | 1,311 (4.8%) |
| Missing | 341 (18.8%) | 715 (39.4%) | 119 (6.6%) | 392 (21.6%) | 246 (13.6%) | 1,813 (6.6%) |
| Education | | | | | | |
| Did not complete high school | 119 (18.6%) | 240 (37.6%) | 53 (8.3%) | 130 (20.3%) | 97 (15.2%) | 639 (2.3%) |
| High school or G.E.D. | 650 (20.3%) | 1,341 (41.9%) | 212 (6.6%) | 682 (21.3%) | 312 (9.8%) | 3,197 (11.7%) |
| Some college | 737 (15.9%) | 1,871 (40.3%) | 349 (7.5%) | 1,093 (23.6%) | 590 (12.7%) | 4,640 (16.9%) |
| Associate's degree | 336 (15.7%) | 924 (43.1%) | 189 (8.8%) | 492 (22.9%) | 205 (9.6%) | 2,146 (7.8%) |
| College graduate | 902 (11.4%) | 2,856 (36.1%) | 644 (8.1%) | 2,111 (26.7%) | 1,408 (17.8%) | 7,921 (28.9%) |
| Post graduate degree | 617 (8.8%) | 2,192 (31.1%) | 605 (8.6%) | 2,101 (29.9%) | 1,523 (21.6%) | 7,038 (25.7%) |
| Missing | 341 (18.8%) | 715 (39.4%) | 119 (6.6%) | 392 (21.6%) | 246 (13.6%) | 1,813 (6.6%) |
| Household Income | | | | | | |
| Under $15,000 | 337 (23.8%) | 564 (39.9%) | 105 (7.4%) | 263 (18.6%) | 144 (10.2%) | 1,413 (5.2%) |
| Between $15,000 and $29,999 | 405 (20.2%) | 871 (43.4%) | 137 (6.8%) | 403 (20.1%) | 191 (9.5%) | 2,007 (7.3%) |
| Between $30,000 and $49,999 | 520 (17.1%) | 1,204 (39.6%) | 264 (8.7%) | 702 (23.1%) | 353 (11.6%) | 3,043 (11.1%) |
| Between $50,000 and $74,999 | 477 (12.9%) | 1,466 (39.6%) | 327 (8.8%) | 916 (24.8%) | 515 (13.9%) | 3,701 (13.5%) |
| Between $75,000 and $99,999 | 347 (10.9%) | 1,202 (37.7%) | 288 (9.0%) | 897 (28.1%) | 458 (14.3%) | 3,192 (11.7%) |
| Between $100,000 and $150,000 | 393 (9.6%) | 1,442 (35.1%) | 321 (7.8%) | 1,154 (28.1%) | 796 (19.4%) | 4,106 (15.0%) |
| Over $150,000 | 271 (7.3%) | 1,066 (28.8%) | 310 (8.4%) | 1,127 (30.4%) | 930 (25.1%) | 3,704 (13.5%) |
| Missing | 952 (15.3%) | 2,324 (37.3%) | 419 (6.7%) | 1,539 (24.7%) | 994 (16.0%) | 6,228 (22.7%) |
| *Survey results among unvaccinated individuals who responded 'yes' or | | | | | | |
| 'not sure' when asked if they would get the SARS-CoV-2 vaccine | | | | | | |

**Table S2** Spatial Lag Regression on Accessibility to SARS-CoV-2 Vaccine Score ($A_{i}$) for each US Census Tract using k-Nearest Neighbor (k=4) Inverse Distance Weight Matrix

|  | **Model 1: Population Density** | |  | **Model 2: Percent Black** | |  | **Model 3: Percent Hispanic** | |  | **Model 4: Age** | |  | **Model 5: Medical Burden‡** | |  | **Model 6: COVID-19 Burden‡** | |
| --- | --- | --- | --- | --- | --- | --- | --- | --- | --- | --- | --- | --- | --- | --- | --- | --- | --- |
|  | **Coefficient** | **[Direct†]** |  | **Coefficient** | **[Direct†]** |  | **Coefficient** | **[Direct†]** |  | **Coefficient** | **[Direct†]** |  | **Coefficient** | **[Direct†]** |  | **Coefficient** | **[Direct†]** |
| **Log of Population Density** | 0.028*** | [0.042] |  | 0.028*** | [0.043] |  | 0.029*** | [0.044] |  | 0.027*** | [0.045] |  | 0.029*** | [0.044] |  | 0.028*** | [0.042] |
| **Percent Black Race** |  |  |  |  |  |  |  |  |  |  |  |  |  |  |  |  |  |
| Q1 (0% – 1.0%] |  |  |  | REF | REF |  |  |  |  |  |  |  |  |  |  |  |  |
| Q2 (1.0% – 4.3%] |  |  |  | 0.006*** | [0.001] |  |  |  |  |  |  |  |  |  |  |  |  |
| Q3 (4.3% – 15.6%] |  |  |  | -0.001*** | [-0.001] |  |  |  |  |  |  |  |  |  |  |  |  |
| Q4 (15.6% – 100%] |  |  |  | -0.003*** | [-0.004] |  |  |  |  |  |  |  |  |  |  |  |  |
| **Percent Hispanic Ethnicity** |  |  |  |  |  |  |  |  |  |  |  |  |  |  |  |  |  |
| Q1 (0% – 2.8%] |  |  |  |  |  |  | REF | REF |  |  |  |  |  |  |  |  |  |
| Q2 (2.8% – 7.6%] |  |  |  |  |  |  | <0.001 | [<0.001] |  |  |  |  |  |  |  |  |  |
| Q3 (7.6% – 20.8%] |  |  |  |  |  |  | -0.010*** | [-0.015] |  |  |  |  |  |  |  |  |  |
| Q4 (20.8% – 100%] |  |  |  |  |  |  | -0.015*** | [-0.023] |  |  |  |  |  |  |  |  |  |
| **Median Age** |  |  |  |  |  |  |  |  |  | -0.0003*** | [-0.0005] |  |  |  |  |  |  |
| **Medical Burden** |  |  |  |  |  |  |  |  |  |  |  |  | 0.060*** | [0.093] |  |  |  |
| **COVID-19 Burden** |  |  |  |  |  |  |  |  |  |  |  |  |  |  |  | -0.462* | [-0.703] |
| **Spatial Lag (ρ)** | 0.892*** | |  | 0.892*** | |  | 0.892*** | |  | 0.893*** | |  | 0.894*** | |  | 0.891*** | |
| **N** | 70027 | |  | 70027 | |  | 70027 | |  | 70027 | |  | 70027 | |  | 70027 | |
| **AIC** | -63879 | |  | -64027 | |  | -64070 | |  | -63864 | |  | -63981 | |  | -63956 | |
| *p < 0.05, **p < 0.01, ***p < 0.001 | | | | | | | | | | | | | | | | | |
| ^†^Direct impact can be interpreted similar to traditional OLS regression coefficient. | | | | | | | | | | | | | | | | | |
| ^‡^Proportion of survey respondents in each census tract who reported COVID-19 related pre-existing conditions (medical burden) or previous contact | | | | | | | | | | | | | | | | | |
| with COVID-19 positive individual (COVID-19 burden). | | | | | | | | | | | | | | | | | |
